# Multi-ethnic polygenic risk modifies the association between *APOL1* high risk genotypes and chronic kidney disease

**DOI:** 10.1101/2021.11.12.21266238

**Authors:** Ha My T. Vy, Faris F. Gulamali, Benjamin S Glicksberg, Orlando Gutierrez, Richard Cooper, Erwin P Bottinger, Judy Cho, Ruth J.F. Loos, Carol R. Horowitz, Ron Do, Girish N Nadkarni

## Abstract

The burden of advanced chronic kidney disease (CKD) falls disproportionately on minorities including African Americans (AAs) and Hispanic Americans (HAs) with admixed ancestry. Even though APOL1 high-risk genotypes increase risk of kidney disease, their penetrance is incomplete, indicating that the modification of APOL1 high risk may be polygenic. For this study, we used three multi-ethnic cohorts with APOL1 high risk genotypes and calculated a multi-ethnic PRS using publicly available summary statistics. We show that CKD risk is significantly modified by a multi-ethnic polygenic risk score. Standardizing population screening for CKD by including APOL1 high-risk genotypes and polygenic risk score may improve risk stratification and outcomes.

---

The burden of advanced chronic kidney disease (CKD) falls disproportionately on minorities including African Americans (AAs) and Hispanic Americans (HAs) with admixed ancestry. The G1/G2 variants at Apolipoprotein L1 (*APOL1*) locus, which confer risk of kidney disease, were subject to positive selection in Africa: *APOL1* high-risk genotypes (G1/G1, G2/G2, or G1/G2) are common in AAs (14-16%) and are present in Afro-Caribbean HAs (∼4%).^1^

Even though *APOL1* high-risk genotypes increase risk of kidney disease, their penetrance is incomplete i.e., only some individuals with high-risk genotypes develop kidney disease. This indicates that alongside interactions with social determinants, common single nucleotide polymorphisms (SNPs) may attenuate the effect of APOL1’s high risk genotypes on kidney disease via epistasis. Prior studies have only shown a small number of epistatic SNPs with small effect sizes, indicating that the modification of *APOL1* high risk may be polygenic.^2^

A polygenic risk score (PRS) aggregates the cumulative effects of millions of common variants across the genome and modifies the effect of monogenic mutations.^3^However, as most genome-wide association studies have been performed in European populations, the available PRS may poorly transfer into diverse populations.^4^ Fortunately, recently multi-ethnic polygenic risk score calculations for diverse populations have been developed. We hypothesized that association between *APOL1* high risk genotypes and CKD is modified by multi-ethnic polygenic background.

For this study, we used three cohorts with *APOL1* high risk genotypes: self-identified Black (n = 890) and Hispanic (n = 151) individuals from Bio*Me* biobank, and Black individuals (n = 1,366) from the UK Biobank^5^ **(Supplemental Table 1)**.

There were 2,407 individuals with *APOL1* high risk genotypes. Mean age ranged from 52-63 years and 50-66% were female. Systolic blood pressure at baseline ranged from 128-140 mm Hg and 18-29% had a history of type 2 diabetes. The proportion of CKD stage 3 ranged from 15 to 30%.

We calculated a multi-ethnic PRS using publicly available summary statistics^6^(**Supplemental Methods**). Using a generalized linear model, we assessed the association between PRS and risk of CKD, adjusting for age, sex, diabetes, and the first ten genetic principal components (**Supplemental Table 2**). In AAs of Bio*Me*, every standard deviation (SD) of PRS was associated with a 13% increase in CKD. Similarly, in the Black British of UKBB, there was a 15% increment in aOR per SD. In contrast, HAs of Bio*Me* cohort had a larger increment in aOR at 63% per SD.

We then conducted a joint meta-analysis of all three groups. The joint meta-analysis in 2407 individuals demonstrated that the increment in odds of CKD per SD PRS was 20% (aOR 1.20, 95% CI 1.06-1.34; p<0.001) (**Figure 1A, Supplemental Table 2**).

**Figure 1.**
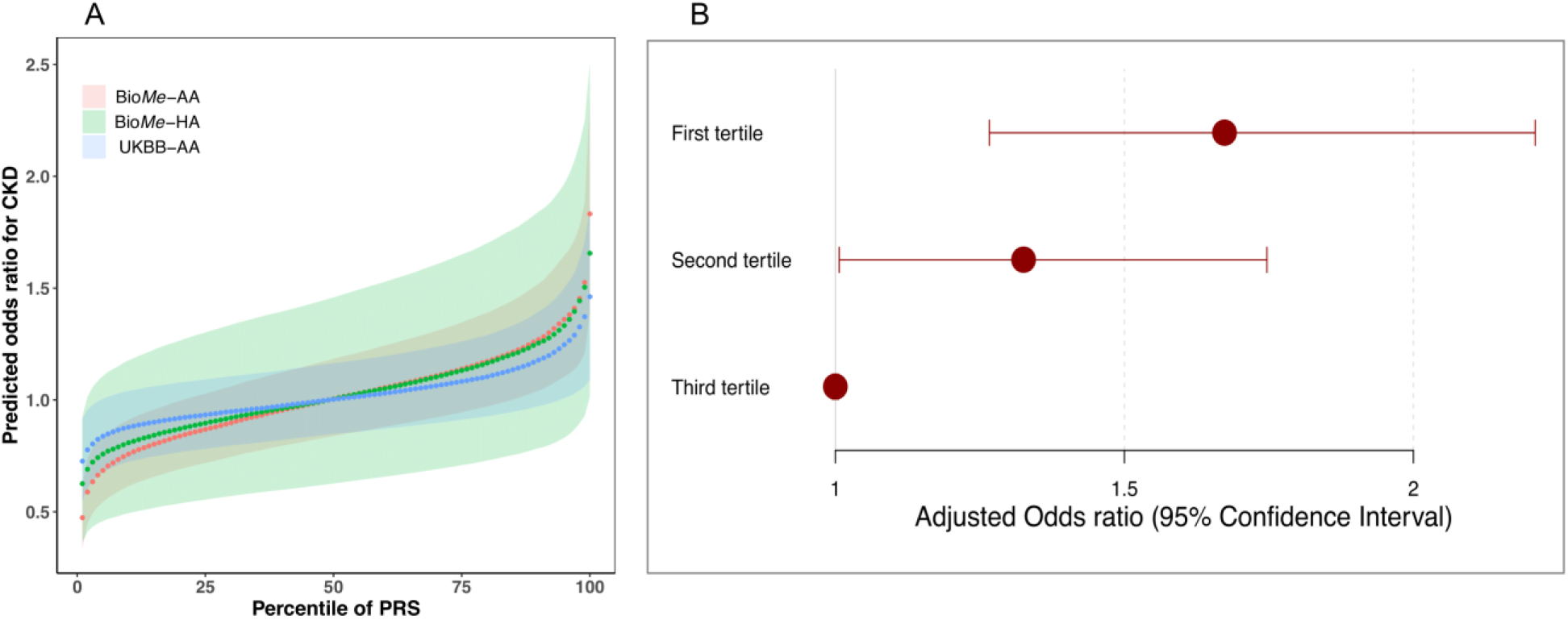
Risk of CKD in diverse patients with *APOL1* high risk genotypes by genome wide polygenic risk continuously (A) and by tertile (B) A. The predicted odds of chronic kidney disease stage 3 or higher by multiethnic polygenic risk continuously. B. The predicted odds of chronic kidney disease stage 3 or higher by tertile of multiethnic polygenic risk by tertile with the reference group being the first tertile.

Next, we stratified carriers and non-carriers in the meta-analysis cohort into three distinct groups by PRS tertile: low (0-33%), intermediate (>33-66%) and high-risk groups (>66-100%). In every individual cohort, the intermediate and high-risk groups had higher odds of CKD compared to the low-risk group in a graded manner (Supplemental Table 2). On meta-analysis, the intermediate risk group had 33% higher adjusted odds (aOR 1.33; 95% CI 1.01-1.75; p=0.04) and the high-risk group had 67% higher adjusted odds (aOR 1.67;95% CI 1.27-2.21; p=0.0003) compared to the low-risk group (**Figure 1B**).

In summary, in three cohorts consisting of 2,407 individuals with high risk *APOL1* genotypes, we show that CKD risk is significantly modified by a multi-ethnic polygenic risk score. *APOL1* is one of the rare mutations which is common and confers risk of serious, chronic disease. As an upstream protein, *APOL1* plays a large multi-modal role in the progression in pore formation, cellular injury, and programmatic cell death, which makes it a prime candidate for epistatic interactions. Here, we show for the first time that a multi-ethnic PRS modifies risk associations.

This work has clinical implications. Genetic screening is becoming increasingly prevalent. Within this genetically high-risk group, further stratification is needed to precisely target resources. For example, individuals with high risk *APOL1* genotypes and high polygenic risk may benefit from early screening followed by targeted interventions to prevent further kidney decline. Standardizing population screening for CKD by including *APOL1* high-risk genotypes and polygenic risk score may improve risk stratification and outcomes. Inclusion of polygenic risk with *APOL1* risk may also allow for more optimal kidney allocation. Finally, PRS may serve as another feature in comprehensive models for risk prediction including baseline Mendelian genetics, biomarkers, clinical and environmental features to power precision medicine approaches to address health disparities in kidney disease.^7^ Limitations of this study include variable comorbidities and outcome rates in the different cohorts. Further efforts are needed to understand how best to disclose integrated risk assessments to patients and clinicians.

## Data Availability

All data produced in the present study are available upon reasonable request to the authors.

---

**Supplemental Table 1.**
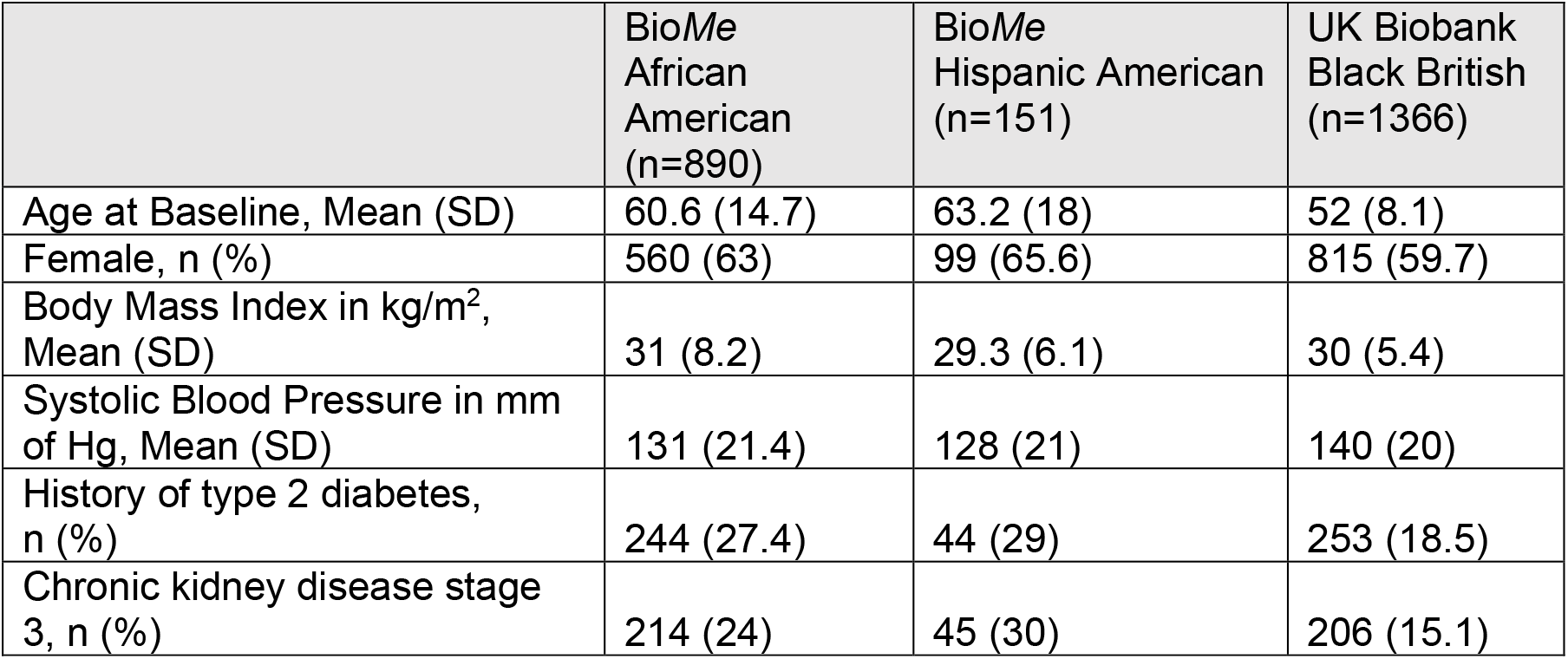
Baseline statistics for the study population (n=2407)

|  | BioMe<br>African<br>American<br>(n=890) | BioMe<br>Hispanic American<br>(n=151) | UK Biobank<br>Black British<br>(n=1366) |
| --- | --- | --- | --- |
| Age at Baseline, Mean (SD) | 60.6 (14.7) | 63.2 (18) | 52 (8.1) |
| Female, n (%) | 560 (63) | 99 (65.6) | 815 (59.7) |
| Body Mass Index in kg/m<sup>2</sup>,<br>Mean (SD) | 31 (8.2) | 29.3 (6.1) | 30 (5.4) |
| Systolic Blood Pressure in mm<br>of Hg, Mean (SD) | 131 (21.4) | 128 (21) | 140 (20) |
| History of type 2 diabetes,<br>n (%) | 244 (27.4) | 44 (29) | 253 (18.5) |
| Chronic kidney disease stage<br>3, n (%) | 214 (24) | 45 (30) | 206 (15.1) |

**Supplemental Table 2.**
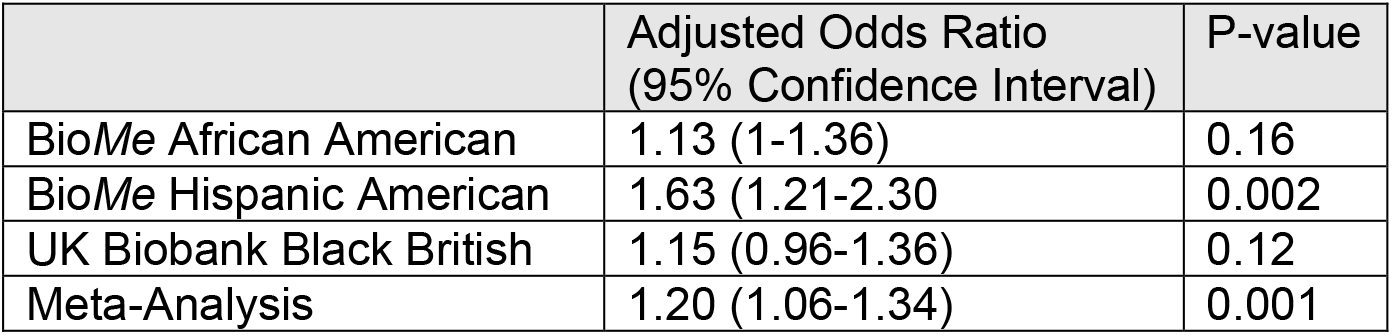
Association tests for kidney disease in diverse individuals with APOL1 high risk genotypes. A generalized linear model was run with CKD Stage 3 or higher as outcome and adjusted for age, sex, type 2 diabetes and ten genetic principal components. Adjusted odds-ratio is per SD increment of multi-ethnic PRS.

|  | Adjusted Odds Ratio<br>(95% Confidence Interval) | P-value |
| --- | --- | --- |
| BioMe African American | 1.13 (1-1.36) | 0.16 |
| BioMe Hispanic American | 1.63 (1.21-2.30) | 0.002 |
| UK Biobank Black British | 1.15 (0.96-1.36) | 0.12 |
| Meta-Analysis | 1.20 (1.06-1.34) | 0.001 |

**Supplemental Table 3.** Adjusted odds of kidney disease in APOL1 high risk genotypes stratified by tertile of multi-ethnic PRS.

| Cohort | PRS Group | Adjusted Odds Ratio<br>(95% Confidence Interval) | P-value |
| --- | --- | --- | --- |
| BioMe African American | First Tertile | 1 (Ref) |  |
|  | Second Tertile | 1.28 (0.84-1.95) | 0.25 |
|  | Third Tertile | 1.48 (1-2.27) | 0.07 |
| BioMe Hispanic American | First Tertile | 1 (Ref) |  |
|  | Second Tertile | 2.90 (0.7-14.4) | 0.16 |
|  | Third Tertile | 8.21 (2.52-32.1) | 0.001 |
| UK Biobank Black British | First Tertile | 1 (Ref) |  |
|  | Second Tertile | 1.30 (0.9-1.9) | 0.17 |
|  | Third Tertile | 1.59 (1.1-2.35) | 0.01 |
| Meta-Analysis | First Tertile | 1 (Ref) |  |
|  | Second Tertile | 1.33 (1.01-1.75) | 0.04 |
|  | Third Tertile | 1.67 (1.27-2.21) | 0.0003 |

## Supplemental Methods

### Study Population

The BioMe population is a retrospective cohort of ∼30,000 individuals with genotyping data that are consented and enrolled from the New York City area. The BioMe biobank is a multi-ethnic cohort, with three main ethnicity categories: African American (N=7381), European American (N=8952), and Hispanic Latin American (N=10,984). CKD was determined by a previously validated phenotyping algorithm.^1^

We included 9,010 individuals who self-reported as Black British in the UK Biobank. CKD was defined by either an ICD code, self-reported, or with CKD-EPI estimated glomerular rate <60 ml/min.

### Multi-ethnic Polygenic Risk Score

Polygenic risk score was calculated by using the effect sizes of a single nucleotide polymorphism-level genome-wide association study, and summing the effect sizes across SNPs within the genome of each individual (*PRS*_*j*_ *=* Σ_*i*_*g*_*ij*_*β*_*i*_ where *g*_*ij*_: the count of effective allele *i* for individual *j*). Multi-ethnic PRS combines polygenic score based on large sample size European training data 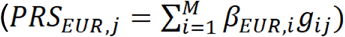 and polygenic score based on training data from the target population 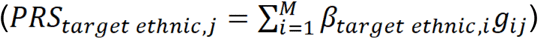 to form a joint estimated PRS 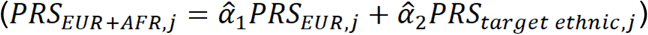. Multi-ethnic polygenic risk scores have been shown to improve risk prediction in diverse populations.

### Genotyping data in BioMe and UKBB

Global Screening Array data is available for N=32,595 participants in BioMe biobank. Sample level quality control was performed and removed individuals with a ethnicity-specific heterozygosity rate that surpassed +/- six standard deviations of the population-specific mean; individuals with a call rate of ≤95%; individuals exhibiting persistent discordance between EMR recorded and genetic sex; and individuals with phenotypically indeterminate sex. For variant level quality control, sites with a call rate below 95% and sites that significantly violate Hardy-Weinberg equilibrium (HWE) when calculated stratified by ancestry (p < 1×10^−5^ in African American and European American, and p < 1×10^−13^ in Hispanic American) were excluded. This resulted in total of 31,911 individuals and 604,869 sites for downstream analysis. Imputation of variants was then performed with Michigan imputation server pipeline using TOPMed reference panel, which yielded genetic information for >184 millions imputed variants.

For our analysis, we included 6942 African American and 10,491 Hispanic American from the sample. In each ethnic group, individuals with relatedness up to 2^nd^degree (estimated by KING)^2^ were excluded using an algorithm that remove one of each related pair while maximizing remaining sample size. This resulted in 6,456 unrelated African American and 9389 unrelated Hispanic American.

The UK Biobank consists of genotype, phenotype, and demographic data of more than 500,000 individuals recruited across the United Kingdom. Individual genotypes were generated from either the Affymetrix Axiom UK Biobank array (∼450,000 individuals) or the UK BiLEVE array (∼50,000 individuals), each contains ∼0.9 million markers.

Additional variants were then imputed using the Haplotype Reference Consortium (HRC) combined with the UK10K haplotype resource, with a total of ∼96 million variants available in the latest released imputed data (version 3). From 9,010 individuals of African ancestry in the data set, we excluded samples that were outliers in heterozygosity or missing rates, samples with putative sex chromosome aneuploidy and one of each pair of samples with relatedness up to the third degree, which lead to 8,993 remaining for subsequent analysis.^3^

### Determination of *APOL1* high risk

*APOL1* high-risk genotypes are defined from two kidney disease risk variants G1 and G2 located in *APOL1* gene. G1 consists of two missense variants at two SNPs in LD: rs73885319 A>G and rs60910145 T>G; and G2 is a 6-bp deletion at SNP rs71785313 TTATAA>-. Since rs71785313 is not available in the genotyping data set, we used the highly linked SNP rs12106505 A>T instead (r^2^ = 0.82).^4^ We defined individuals that were homozygous for either G1 or G2 or carries at least one G1 copy and one G2 copy (G1/G1; G2/G2; or G1/G2) is defined as high-risk genotype.

### Statistical Analysis

We included key covariate variables specifically age, sex, and Type 2 Diabetes. We aggregated covariate statistics by ethnicity and computed means with standard deviations. We fit a generalized linear model, regressing CKD on polygenic risk score, *APOL1* high risk genotypes and the covariates and calculated the best linear unbiased estimator. We reported odds ratios and their corresponding p-values.

## Notes

### Competing Interest Statement

The authors have declared no competing interest.

### Funding Statement

The study was funded by R01DK127139.

### Author Declarations

Ethics committee/IRB of Icahn School of Medicine at Mount Sinai Hospital gave ethical approval for this work.

## References

1. Nadkarni GN, Gignoux CR, Sorokin EP, Daya M, Rahman R, Barnes KC, Wassel CL, Kenny EE. Worldwide Frequencies of APOL1 Renal Risk Variants. N Engl J Med. 2018 Dec 27;379(26):2571-2572. PMID: 30586505; PMCID: PMC6482949.

2. Langefeld CD, Comeau ME, Ng MCY, Guan M, Dimitrov L, Mudgal P, Spainhour MH, Julian BA, Edberg JC, Croker JA, Divers J, Hicks PJ, Bowden DW, Chan GC, Ma L, Palmer ND, Kimberly RP, Freedman BI. Genome-wide association studies suggest that APOL1-environment interactions more likely trigger kidney disease in African Americans with nondiabetic nephropathy than strong APOL1-second gene interactions. Kidney Int. 2018 Sep;94(3):599-607. Epub 2018 Jun 7. PMID: 29885931; PMCID: PMC6109415.

3. Fahed AC, Wang M, Homburger JR, Patel AP, Bick AG, Neben CL, Lai C, Brockman D, Philippakis A, Ellinor PT, Cassa CA, Lebo M, Ng K, Lander ES, Zhou AY, Kathiresan S, Khera AV. Polygenic background modifies penetrance of monogenic variants for tier 1 genomic conditions. Nat Commun. 2020 Aug 20;11(1):3635. PMID: 32820175; PMCID: PMC7441381.

4. Márquez-Luna C, Loh PR; South Asian Type 2 Diabetes (SAT2D) Consortium; SIGMA Type 2 Diabetes Consortium, Price AL. Multiethnic polygenic risk scores improve risk prediction in diverse populations. Genet Epidemiol. 2017 Dec;41(8):811-823. Epub 2017 Nov 7. PMID: 29110330; PMCID: PMC5726434.

5. Bycroft C, Freeman C, Petkova D, Band G, Elliott LT, Sharp K, Motyer A, Vukcevic D, Delaneau O, O’Connell J, Cortes A, Welsh S, Young A, Effingham M, McVean G, Leslie S, Allen N, Donnelly P, Marchini J. The UK Biobank resource with deep phenotyping and genomic data. Nature. 2018 Oct;562(7726):203–209. doi: 10.1038/s41586-018-0579-z. Epub 2018 Oct 10. PMID: 30305743; PMCID: PMC6786975.

6. Pattaro C, Teumer A, Gorski M, et al. Genetic Associations at 53 Loci Highlight Cell Types and Biologic Pathways for Kidney Function. Nat Commun. 2016 Jan 21; 7:10023

7. Paranjpe I, Chaudhary K, Paranjpe M, O’Hagan R, Manna S, Jaladanki S, Kapoor A, Horowitz C, DeFelice N, Cooper R, Glicksberg B, Bottinger EP, Just AC, Nadkarni GN. Association of APOL1 Risk Genotype and Air Pollution for Kidney Disease. Clin J Am Soc Nephrol. 2020 Mar 6;15(3):401-403. Epub 2020 Feb 20. PMID: 32079610; PMCID: PMC7057301.

## References

1. Nadkarni GN, Gottesman O, Linneman JG, Chase H, Berg RL, Farouk S, Nadukuru R, Lotay V, Ellis S, Hripcsak G, Peissig P, Weng C, Bottinger EP. Development and validation of an electronic phenotyping algorithm for chronic kidney disease. AMIA Annu Symp Proc. 2014 Nov 14;2014:907-16. PMID: 25954398; PMCID: PMC4419875.

2. Manichaikul A, Mychaleckyj JC, Rich SS, Daly K, Sale M, Chen WM. Robust relationship inference in genome-wide association studies. Bioinformatics. 2010 Nov 15;26(22):2867–73. doi: 10.1093/bioinformatics/btq559. Epub 2010 Oct 5. PMID: 20926424; PMCID: PMC3025716.

3. Bycroft C, Freeman C, Petkova D, Band G, Elliott LT, Sharp K, Motyer A, Vukcevic D, Delaneau O, O’Connell J, Cortes A, Welsh S, Young A, Effingham M, McVean G, Leslie S, Allen N, Donnelly P, Marchini J. The UK Biobank resource with deep phenotyping and genomic data. Nature. 2018 Oct;562(7726):203–209. doi: 10.1038/s41586-018-0579-z. Epub 2018 Oct 10. PMID: 30305743; PMCID: PMC6786975.

4. Genovese G, Friedman DJ, Pollak MR. APOL1 variants and kidney disease in people of recent African ancestry. Nat Rev Nephrol. 2013 Apr;9(4):240–4. doi: 10.1038/nrneph.2013.34. Epub 2013 Feb 26. PMID: 23438974.

